# A coupled model to value indirect impacts of maternal, newborn and child health interventions

**DOI:** 10.1101/2025.11.06.25339740

**Authors:** Tom Walsh, Joshua P. Vogel, Maureen Makama, Romesh Abeysuriya, Nick Scott

## Abstract

**Background:** Outcomes associated with maternal, newborn and child health (MNCH) conditions have improved significantly over the past 20 years, however, they are not on track to meet the global SDG targets by 2030. Emerging interventions have the potential to greatly increase access to care, but estimating their impacts is difficult due to complicated interactions between interventions. Models exist to quantify outcomes and costs associated with scaling up interventions related to MNCH (e.g., The Lives saved tool (LiST)), but none have been designed to flexibly integrate novel interventions. This study aimed to develop a model that can estimate the impact of standard-of-care (SOC) and novel interventions on MNCH conditions, and present an example use case.

**Methods:** We developed a compartment-based model with 15 interconnected population- and condition-specific modules for pregnant women (in different trimesters), newborns and children <5 years to quantify impacts of bespoke novel interventions. The model included modules for anemia, pre-eclampsia, obstructed labour, hemorrhage, maternal sepsis, preterm, small-for-gestation-age (SGA), respiratory distress syndrome (RDS), neonatal sepsis, birth asphyxia, wasting (neonates and children), stunting and modules that captures neonatal and childhood conditions not captured elsewhere in the model.range of conditions related to MNCH conditions.

**Results:** The model captures dynamic interactions between MNCH conditions, including across trimesters, and accounts for the full impact of early detection and prevention of conditions during pregnancy. In an example use case, the model identified that scaling-up standard-of-care and novel diagnostic interventions for anemia and pre-eclampsia plus AI-ultrasound (for early detection of obstructed labour, hemorrhage and birth outcome risks, leading to facility referrals) to 60% could avert 207 million DALYs globally over 2024-2040.

**Conclusions:** We have developed a modeling tool which can be flexibly adapted to assess the impacts of novel interventions for MNCH conditions, and can be applied at a global, regional, national or sub-national level.

**Funding:** Bill and Melinda Gates Foundation

## Introduction

Each day there are approximately 500 maternal deaths and 5,000 neonatal deaths globally(1). Approximately 95% of these deaths occur in low-and middle-income countries (LMICs), and most are a result of preventable or treatable conditions. In 2015, the Sustainable Development Goals (SDG) initiative set targets for neonatal and maternal mortality to be reduced to below 12 per 1,000 live births and 70 per 100,000 live births by 2030, respectively(2). While maternal, newborn and child health (MNCH) outcomes have drastically improved throughout much of the world over the last two decades, progress in many LMICs stalled during the COVID-19 pandemic. Significant acceleration of progress will be required to reach global and national SDG targets for maternal and newborn deaths by 2030.

Mathematical models can simulate interactions between MNCH conditions. They are a useful tool to estimate the relative benefits of scaling up different packages of interventions. Models already exist which quantify the impact of interventions on MNCH conditions - the most widely used being the lives saved tool (LiST)(3). LiST is an industry standard modelling tool that has been used in many analyses to inform public policy surrounding investment into MNCH. However, there are unique challenges associated with modelling MNCH interventions that limit the utility of existing tools.

MNCH conditions have complex interactions, such that preventing or treating one condition can often have impacts on the incidence of other conditions. For example, maternal anemia is a risk factor for maternal mortality(4, 5), being small-for-gestational-age (SGA) (6) and for being born preterm(7). SGA and preterm birth are themselves risk factors for neonatal mortality(8), stunting and wasting(9). Therefore, population-level initiatives to prevent anaemia could reduce the prevalence of multiple other ‘downstream’ conditions. These dynamics are important to include, in order to capture the full direct and downstream benefits of introducing MNCH interventions.

An additional complexity is that to effectively treat or manage many of these conditions, diagnosis and clinical monitoring interventions are required. These may not always be available, particularly in LMICs. For example, the upfront cost of antenatal ultrasound devices, as well as shortages of healthcare professionals who can use them accurately, prevents conditions such as fetal growth restriction from being detected and hence limits the impact that subsequent interventions can have. Similarly, pre-eclampsia is a leading cause of maternal mortality, yet diagnosis relies on accurate measurement of blood pressure, proteinuria and other forms of organ dysfunction, which may not always be available. In the case of pre-eclampsia, there is a need for new risk screening and diagnostic tools to expand coverage, and there are pipelines of such products coming to market(10). Quantifying the impacts of scaling up current MNCH interventions, and what additional value novel interventions may bring, helps to prioritize investments in health and research programs. While some other models, including LiST, capture interactions between interventions, risk factors and outcomes, they are not able to flexibly integrate new interventions, or consider more complex interactions.

To address this, we developed a model in collaboration with the Bill and Melinda Gates Foundation (BMGF), which captures the complex interactions between interventions, risk factors and outcomes and also allows for flexible integration of new interventions. The model enables scenarios to be projected, where combinations of emerging technologies and existing interventions are implemented and scaled up. This allows the user to identify packages with the greatest potential for impact. In this paper, we describe the key aspects of the model design and demonstrate an example use case.

## Methods

The model outlined in this study was developed around a set of interventions included in a study commissioned by and conducted for the BMGF. This analysis implements a subset of these interventions. The interventions under consideration in the BMGF study informed the structure and guided the model decisions.

### Model overview

Fifteen distinct, interconnected MNCH modules were constructed to account for the intergenerational linkages between maternal, fetal, newborn and infant risk factors and outcomes (Figure 1, Table 1). Each module is a deterministic compartmental model, which is linked to one or more other modules through shared parameters (e.g. the prevalence of maternal anemia is an output of the anemia module, and an input parameter for the birth outcome module).

**Figure 1.**
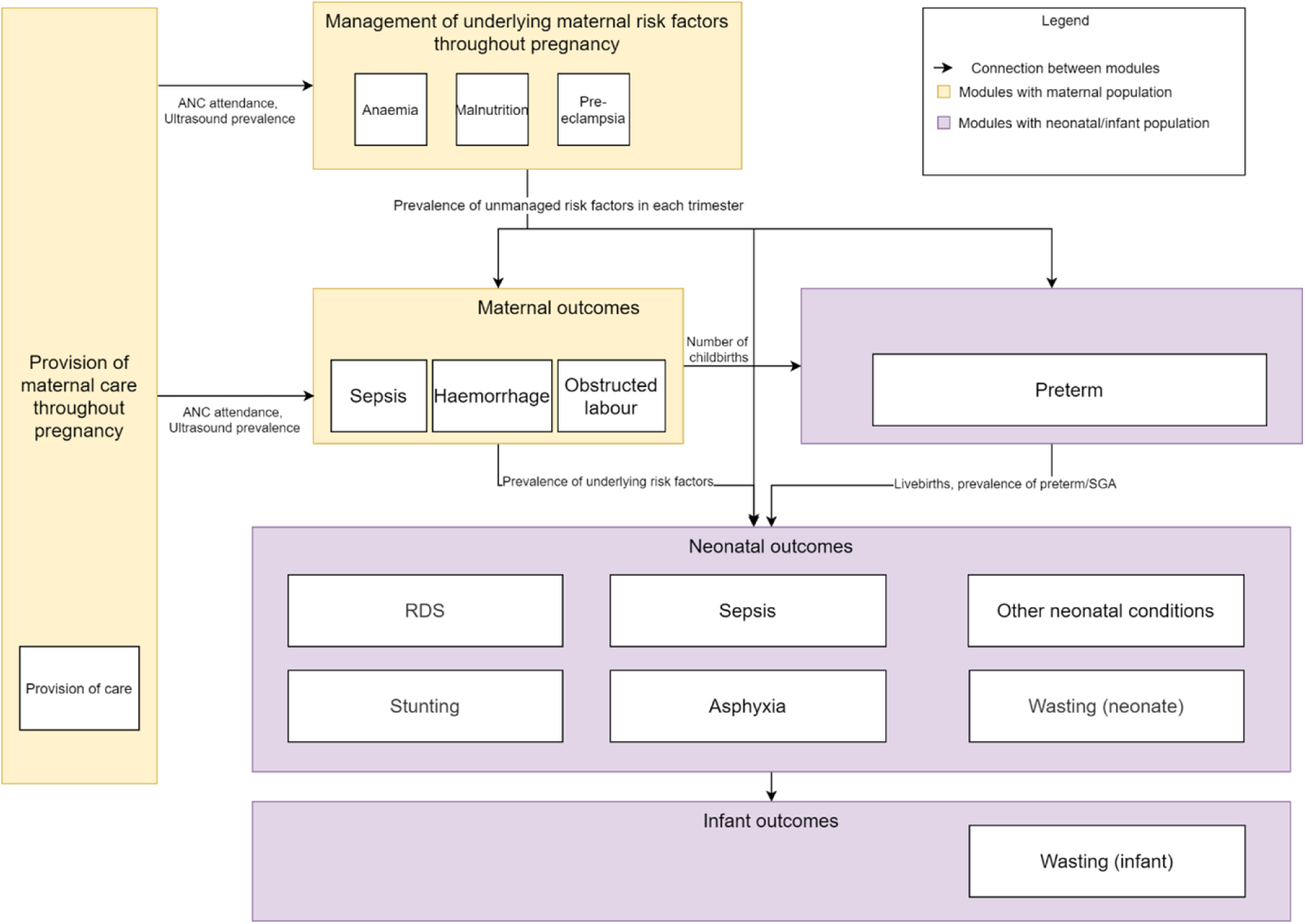
Model structure for capturing morbidities and mortalities for specific conditions, highlighting interactions between conditions.

**Table 1.** Key aspects of modules within the framework.

| **Timestep** | **Module** | **Description** | **Inputs from other modules** | **Outputs for a given intervention scenario** |
| --- | --- | --- | --- | --- |
| Pregnancy<br>(three distinct trimesters modelled) | ANC | Tracks ANC attendance through pregnancy and allows ultrasound coverage to be scaled up<br>Tracks referral facility attendance for childbirth. Referral facility attendance is a requirement for a number of childbirth interventions and reduces the impact of conditions that occur at childbirth. | Number of pregnant women | Proportion attending ANC<br>Referral facility<br>Ultrasound coverage<br>Number of pregnant women that continue to childbirth |
|  | Anemia<br>(Intrapartum and postpartum) | Tracks anemia status in pregnant women, split into mild, moderate and severe. | Number of pregnant women<br>Proportion attending ANC<br>Hemorrhage prevalence (postpartum anemia) | Number of pregnant women with anemia (3rd trimester and postpartum)<br>Number of DALYs due to anemia (during pregnancy and postpartum) |
|  | Pre-eclampsia | Tracks preeclampsia in pregnant women with early onset and late onset pre-eclampsia and the impact of screening and treatment interventions. | Number of pregnant women<br>Proportion attending ANC | Number of morbidities, mortalities and DALYs due to pre-eclampsia (for each trimester)<br>Prevalence of early and late onset pre-eclampsia |
| Childbirth | Obstructed labour | Tracks obstructed labour in childbirth. Obstructed labour can be averted by a medically necessary Caesarean section. | Number of pregnant women that continue to childbirth<br>Proportion with RF delivery<br>Ultrasound coverage (Necessary c-section identification) | Number of morbidities, mortalities and DALYs due to obstructed labour<br>Prevalence of obstructed labour (with and without c-section) |
|  | Postpartum hemorrhage | Tracks postpartum hemorrhage in pregnant women during the modelled childbirth timestep. | Number of pregnant women that continue to childbirth<br>Proportion with RF delivery<br>Ultrasound coverage (identification of likely hemorrhage for referral to higher level facility) | Number of morbidities, mortalities and DALYs due to hemorrhage |
|  |  |  | Anemia prevalence |  |
|  | Maternal nutrition interventions | Tracks coverage of maternal nutrition interventions in pregnant women. | Number of pregnant women that continue to childbirth | Coverage of maternal nutrition interventions (ie, MMS, IFAS) |
|  | Maternal sepsis | Tracks sepsis in women during childbirth. | Number of pregnant women that continue to childbirth | Number of morbidities, mortalities and DALYs due to maternal sepsis<br>Prevalence of maternal sepsis |
|  | Preterm/SGA | Tracks preterm and/or SGA birth outcomes. Associated morbidities can be reduced in severity by delivery at a referral facility. Facility delivery for imminent preterm and/or SGA births can be increased by accurate detection with ultrasound. | Number of pregnant women that continue to childbirth<br>Prevalence of maternal anemia (3rd trimester)<br>Prevalence of preeclampsia | Number of morbidities, mortalities and DALYs due to preterm and/or SGA<br>Prevalence of preterm and/or SGA |
| Neonatal time period | RDS | Tracks birth asphyxia and subsequent neonatal encephalopathy in neonates during the neonatal period and calculates number of births which are stillbirth during pregnancy. | Number of pregnant women that continue to childbirth<br>Prevalence of obstructed labour<br>Prevalence of obstructed labour without c-section<br>Prevalence of maternal sepsis<br>Coverage of maternal nutrition interventions | Number of live births<br>Number of morbidities, mortalities and DALYs due to neonatal encephalopathy<br>Number of and DALYs due to stillbirth |
|  | RDS | Tracks RDS in preterm neonates during the neonatal period. Any prevention can only be targeted towards those who are identified as preterm. | Number of live births<br>Prevalence of preterm births | Number of morbidities, mortalities and DALYs due to RDS |
|  | Neonatal sepsis | Tracks neonatal sepsis in neonates during the neonatal period. | Number of live births<br>Prevalence of preterm births | Number of morbidities, mortalities and DALYs due to neonatal sepsis |
|  | Stunting | Tracks stunting risk during the neonatal time period, and associated disability due to stunting. | Number of live births<br>Coverage of maternal nutrition interventions<br>Prevalence of preterm/SGA birth outcomes | Number of morbidities, mortalities and DALYs due to stunting |
|  | Other neonatal conditions (encephalitis, meningitis, enteric infections, respiratory infections, and malaria) | Tracks neonatal conditions otherwise not captured in the model during the neonatal period. | Number of live births | Number of morbidities, mortalities and DALYs due to other neonatal conditions |
| Infant - childhood time period | Wasting | Tracks wasting in infants and children from 8 days to 5 years of life. Each cohort of neonates in the 3 month time period are tracked for the 5 years with the timesteps split into 8 days - 5 months, 6 months - 18 months and 19 months to 59 months. | Number of surviving neonates<br>Prevalence of preterm/SGA birth outcomes | Number of morbidities, mortalities and DALYs due to wasting |

Women enter the model population when they become pregnant and are tracked across trimesters, with childbirth and immediate postpartum outcomes recorded. Within each trimester, fetal development is also modelled. The model captures the prevalence of different birth outcomes (pre-term and/or small for gestational age [SGA]) and tracks newborn and infant outcomes up to age five. By explicitly modelling the three trimesters for pregnant women, the model can account for the impact of early intervention during pregnancy.

The model produces outputs for the prevalence of a range of MNCH outcomes, morbidities and mortality, which are converted to disability-adjusted life years (DALYs). The 15 modules are split into 6 distinct sections as shown in Figure 1 and as described below.

#### Provision of care

The provision of care module tracks the coverage of antenatal care during each trimester, which influences the maximum achievable coverage of other interventions, as well as referrals to differently equipped facilities for birth (Appendix D).

#### Maternal risk factors

Maternal risk factors that are modelled include anemia, pre-eclampsia and undernutrition. The maternal anemia and pre-eclampsia modules track the development, detection and management of these conditions across trimesters, while the maternal nutrition module tracks the prevalence and treatment of undernutrition during pregnancy. Outputs of these modules are the prevalence of maternal anemia (mild, moderate or severe), prevalence of preeclampsia (treated, untreated), pre-eclampsia-related mortality, and the prevalence of maternal undernutrition. These outputs are themselves inputs for the maternal outcome, birth outcome, and newborn outcome sections.

#### Maternal outcomes

Maternal outcomes include peripartum sepsis, postpartum hemorrhage and obstructed labour occurring around the time of childbirth, and their prevalence is dependent on outputs of the provision of care and maternal risk factor modules. Outputs include the prevalence of morbidities and mortality due to each condition. These outputs are themselves risk factors for the neonatal modules.

#### Birth outcomes

The birth outcome timestep also tracks the prevalence of preterm and SGA births, as well as associated morbidities and mortalities. This module is dependent on outputs from the provision of care and maternal risk factor modules, and the prevalence of preterm and/or SGA birth influence the neonatal outcomes module.

#### Neonatal outcomes

Neonatal outcomes that are modelled include respiratory distress syndrome (RDS), sepsis, stunting, asphyxia, wasting, other neonatal conditions. Other neonatal conditions cover mortalities related to neonatal conditions that are not captured elsewhere in this model. The modules track the development, detection and management of these conditions in the neonatal period (up to 28 days postpartum). Outcomes of these modules are the prevalence of RDS, sepsis, stunting, asphyxia, wasting and other neonatal conditions, and mortality due to RDS, sepsis, neonatal encephalopathy, wasting and other neonatal conditions. These outcomes are themselves inputs for the infant outcome modules.

#### Infant and child outcomes

The infant and child outcome that is modelled is wasting in the infant and childhood time period. The modules track the development, detection and management of wasting in the infant period up to 5 years post birth. Outcomes include morbidities and mortalities due to wasting.

#### Modules

Key aspects of each module within the framework are described in Table 1.

### Model parameters

The model requires setting-specific inputs for demographics, prevalence of MNCH conditions, and maternal and newborn mortality rates. Parameters are also required for estimated relative risks between conditions, as well as baseline intervention and health system coverage. For many settings, the required data can be obtained from the Institute for Health Metrics and Evaluation (IHME)(11) and relative risks are available from systematic reviews and meta-analyses (Appendix A).

### Model outputs

Health benefits that are estimated when the coverage of interventions in the model increases include cases of anemia, PPH, obstructed labour, maternal sepsis, preterm, SGA, neonatal encephalopathy, wasting, stunting, neonatal sepsis averted and maternal deaths, stillbirths and newborn deaths averted.

This model contrasts to previous models in that it aims to explicitly account for intervention impacts via direct intervention impact on specific conditions, and subsequent indirect impacts on other, downstream conditions. In such a way we attempt to both recategorize previously captured impacts and account for previously uncaptured impacts.

### Interventions

Model outputs can be impacted by interventions. To be included in the model, an intervention requires one or more target modules, a target parameter within those modules, effect size on the parameter, baseline coverage, and maximum achievable coverage. For example, standard-of-care (SOC) iron tablets during pregnancy target the anemia module. It has an estimated relative reduction of 95% for severe, 88% for moderate and 76% for mild anemia and a maximum achievable coverage of 95% based on the proportion who attend ANC.

Changes in intervention coverage therefore cause changes in outcomes of the target modules (“direct effects”), as well as changes in downstream outcomes (“indirect effects”). The attributes for an example setting are described in Table 2.

**Table 2.** Summary of direct and indirect impacts of each interventions used in the example application.

| **Intervention** | **Direct impacts** | **Indirect impacts** | **Coverage denominators** | **Effect size (relative risk unless otherwise specified)** | **Data source** |
| --- | --- | --- | --- | --- | --- |
| Ready-to-use therapeutic foods | Reduce wasting |  | Children with wasting (hospitalized or not hospitalized) for 6 to 18 months and 19 to 59 months | Severe: 0.58<br>Moderate: 0.36 | Isanaka et al. JAMA 2009.(13) |
| Iron folic acid tablets | Reduces anemia | Reduces hemorrhage<br>Reduces postpartum anemia | Identified first/second trimester mild/moderate/severe anemia attending ANC | Severe treatment: 0.95<br>Moderate treatment: 0.88<br>Mild treatment: 0.76 | Oh C, et al. Nutrients. 2020(14) |
| Pre-eclampsia urine dipstick test | Detects pre-eclampsia of all severity | Reduces stillbirth | Women who will experience second/third trimester preeclampsia attending ANC | Sensitivity = 0.68 (Value is for women with suspected preeclampsia but is a reasonable estimate for the modelled population.) | Teeuw HM, et al. Diagnostic accuracy of urine dipstick tests for proteinuria in pregnant women suspected of preeclampsia: A systematic review and meta-analysis. Pregnancy Hypertens. 2022 Mar;27:123-130. (15) |
| SOC Ultrasound | Identifies risk factors for obstructed labor<br>Identifies risk factors for hemorrhage<br>Estimates gestational age allowing | Reduces adverse outcomes associated with Hemorrhage, Obstructed labour and Preterm/SGA<br>Reduces stillbirth due to reduction in adverse effects of obstructed labour<br>Reduces neonatal encephalopathy/ RDS | All women ever attending ANC (Maximum 50% coverage) | Identification of risk factors for<br>Obstructed labor: 0.9 (Assuming all cases require c-section)<br>Hemorrhage: 0.57<br>Preterm: 0.5<br>SGA: 0.6 | Estimates from primary data from VITAL Pakistan Pregnancy Cohort. Ultrasound-identifiable risk factor for hemorrhage defined as presence of multiple, placenta previa, or breech presentation (breech is the only type of malpresentation measured in the database but this is the most common type.) |
|  | more accurate identification of preterm birth and SGA | adverse outcomes due to referral facility birth |  |  |  |
| Pre-eclampsia treatment (severe PE only) | Reduces impacts of severe pre-eclampsia | Reduces:<br>• non-RDS preterm<br>• SGA/<br>• RDS<br>• Stillbirth<br>due to reduced pre-eclampsia | Women with identified severe PE attending ANC | Maternal morbidity: 0.6<br>Maternal mortality: 0.6 | Duley et al. Cochrane Database Syst Rev. 2010 (16) |
| Hb test | Diagnoses anemia | Reduces anemia by increasing treatment<br>Reduces postpartum hemorrhage<br>Reduces:<br>• Non- RDS preterm<br>• SGA<br>• Stunting<br>• Wasting<br>due to reduced anemia | All pregnant women (first/second trimester) attending ANC | Sensitivity = 0.667 | Assumed value based on HemoCue system:<br>1. Fothergill et al. J Nutr. 2023 (17)<br>2. Alarcón-Yaquetto et al. PLoS One. 2023(18)<br>3. Sanchis-Gomar et al. J Lab Autom 2013(19) |
| Non-invasive Hb test | Diagnoses anemia | Reduces anemia by increasing treatment<br>Reduces hemorrhage<br>Reduces Non- RDS preterm/SGA/Stunting/ | All pregnant women (first/second trimester) attending ANC | Sensitivity = 0.95 | Assumption based on expert opinion |
|  |  | Wasting due to reduced anemia |  |  |  |
| Pre-eclampsia risk screening and prevention | Identifies and reduces likelihood of pre-eclampsia occurring | Reduces Non- RDS preterm/SGA/ RDS /Stillbirth due to reduced pre-eclampsia | Women at high risk of PE attending first trimester ANC | Early onset PE incidence: 0.8 | Assumption consistent with Duley et al. 2019 Cochrane (18% reduction) |
| AI ultrasound | Identifies risk factors for obstructed labor<br>Identifies risk factors for hemorrhage<br>Estimates gestational age allowing more accurate identification of preterm birth and SGA<br>Increases coverage of ultrasound compared to SOC ultrasound | Reduces adverse outcomes associated with Hemorrhage, Obstructed labour and Preterm/SGA<br>Reduces stillbirth due to reduction in adverse effects of obstructed labour<br>Reduces neonatal encephalopathy/RDS adverse outcomes due to referral facility birth | All women ever attending ANC (note: intervention increases coverage of this population) | Identification of risk factors for<br>• Obstructed labor: 0.9<br>• Hemorrhage: 0.57<br>• Preterm: 0.5<br>• SGA: 0.6<br>Note: main impact is higher coverage for AI ultrasound due to accessibility<br>Note: other impacts are captured through links between conditions | Same values as for SOC ultrasound |

In the model, both standard-of-care and novel interventions modify the prevalence and outcomes of conditions. Interventions can be flexibly introduced and removed on a per-analysis basis.

### Model calibration

The model is calibrated such that when it is run with baseline intervention coverage, it reproduces the burden of maternal anemia, preeclampsia, maternal sepsis, postpartum hemorrhage, obstructed labour, preterm births, SGA births, neonatal deaths (by cause), stunting, wasting and infant deaths (by cause). This is achieved by varying the probability of developing these conditions in each module. It is assumed that a single year captures the relationship between conditions within the model. This calibration occurs with SOC interventions included in the model.

### Example application

We describe an example use case of the model based on a recent study by the BMGF examining a set of standard of care and novel interventions under development(12). We considered two scenarios – a realistic achievable coverage and an aspirational coverage for these interventions. A realistic coverage was defined as 60% of maximal achievable coverage and an aspirational coverage was defined as 90% of maximal achievable coverage.

#### Setting

We ran the model for LMICs, grouped into three aggregated settings: Sub-Saharan Africa, South Asia and other LMICs. The parameters associated with each modelled setting represent the aggregate of LMICs that it contains. These were chosen to align with a previous BMGF study(12).

### Interventions

Interventions included in this analysis were based on the recent BMGF study. Six existing interventions and three emerging interventions (i.e. likely to be available in the near term) were included in the modelling analysis, described in detail in Table 2. Evidence for the impact of these interventions is given in Table 2.

#### Scenarios design

MNCH outcomes were projected across LMICs from 2024-2040 under the following intervention scenarios:

- **Baseline:** Interventions remain at current levels
- **Baseline + individual intervention scale-up:** Individual products scaled up in isolation to reach either 60% or 95% coverage by 2030 (except for SOC Ultrasound which was scaled up to 50%, see Table 2)
- **SOC scale up:** All SOC interventions scaled up together to reach either 60% or 95% by 2030.
- **SOC + novel diagnostics scale up:** All SOC plus novel diagnostics interventions scaled up together to reach either 60% or 95% by 2030.

In cases where the SOC product has baseline coverage greater than 60%, coverage was maintained for the 60% coverage scenarios.

#### Data

Model inputs for demographic, epidemiological, and health system parameters were taken from available primary data sets, IHME GBD 2019 estimates, and published literature (Appendix A). All models used region-specific data inputs where possible for three regional groupings: South Asia; sub-Saharan Africa; and other LMICs comprising countries in Latin America, North Africa/Middle East, and East/Southeast Asia/Oceania. We based intervention effect size assumptions on published literature and available primary data. Coverage parameter values were constrained by intervention delivery channel access (e.g., antenatal care coverage, in-facility delivery coverage).

## Results

### Comparison between individual intervention scenarios

In the baseline scenario, there were an estimated 5.9 billion DALYs from the modelled conditions in LMICs over 2024-2030 (Table 3). Indirect impacts made up the majority of DALYs averted (~93% cumulative in the 60% coverage scenario and ~91% cumulative in the 95% coverage scenario), except for RUTF scale-up which impacts only infants directly in the model. From the individual intervention scenarios, the AI ultrasound had the greatest impact, followed by the SOC ultrasound, due to their large downstream indirect impacts. The AI ultrasound performed better due to higher possible coverage, as it could be performed by lower cadre healthcare workers. For the 95% scenarios, the highest impact interventions in terms of DALY reductions were AI ultrasound, SOC ultrasound and the preeclampsia screening.

**Table 3.** Reduction of total DALYs in each scenario by region compared to the baseline scenario.

| Scenario | Impact type | Total DALYs averted (compared to baseline) |  |
| --- | --- | --- | --- |
|  |  | 60% + individual intervention scale up (Realistic scenarios) | 95% + individual intervention scale up (Aspirational scenarios) |
| *Baseline* | *Total (not averted)* | 6,317,221,237 | 6,317,221,237 |
| Baseline + SOC RUTF | Direct | 0 | 3,156,725 |
|  | Indirect | 0 | 0 |
| Baseline + SOC Iron tablets | Direct | 1,185,059 | 1,798,971 |
|  | Indirect | 7,990,134 | 11,598,604 |
| Baseline + SOC Hb test | Direct | 0 | 32,262 |
|  | Indirect | 0 | 209,850 |
| Baseline + SOC preeclampsia urine dipstick test | Direct | 6,927 | 148,910 |
|  | Indirect | 46,617 | 615,537 |
| Baseline + SOC ultrasound | Direct | 8,195,552 | 8,195,552 |
|  | Indirect | 93,237,748 | 93,237,748 |
| Baseline + SOC preeclampsia treatment | Direct | 0 | 0 |
|  | Indirect | 0 | 654,049 |
| Baseline + non-invasive Hb test | Direct | 168,956 | 192,349 |
|  | Indirect | 1,123,270 | 1,284,958 |
| Baseline + preeclampsia screening and treatment | Direct | 1,182,011 | 1,182,011 |
|  | Indirect | 58,248,758 | 58,248,758 |
| Baseline + AI ultrasound | Direct | 10,831,668 | 20,669,950 |
|  | Indirect | 122,577,369 | 227,395,843 |
| SOC Scale up | Direct + Indirect | 110,578,189 | 121,423,390 |
| SOC + diagnostics scale up | Direct + Indirect | 207,180,195 | 365,294,286 |

In particular, in the 95% scenario, AI ultrasound reduced DALYs attributable to birth asphyxia by 12%, obstructed labour by 11% and hemorrhage by 8%; SOC ultrasound reduced DALYs attributable to birth asphyxia by 5%, obstructed labour by 5%, and hemorrhage by 3%; preeclampsia screening reduced DALYs attributable to preeclampsia by 5%, stillbirth by 3% and non-RDS preterm by 1%.

### Scale up scenarios

Compared to the baseline scenario, scaling up all SOC interventions to 60% or 95% coverage averted an additional **111 million and 121 million DALYs** over 2024-2040, respectively (Table 3, Figure 2).

**Figure 2.**
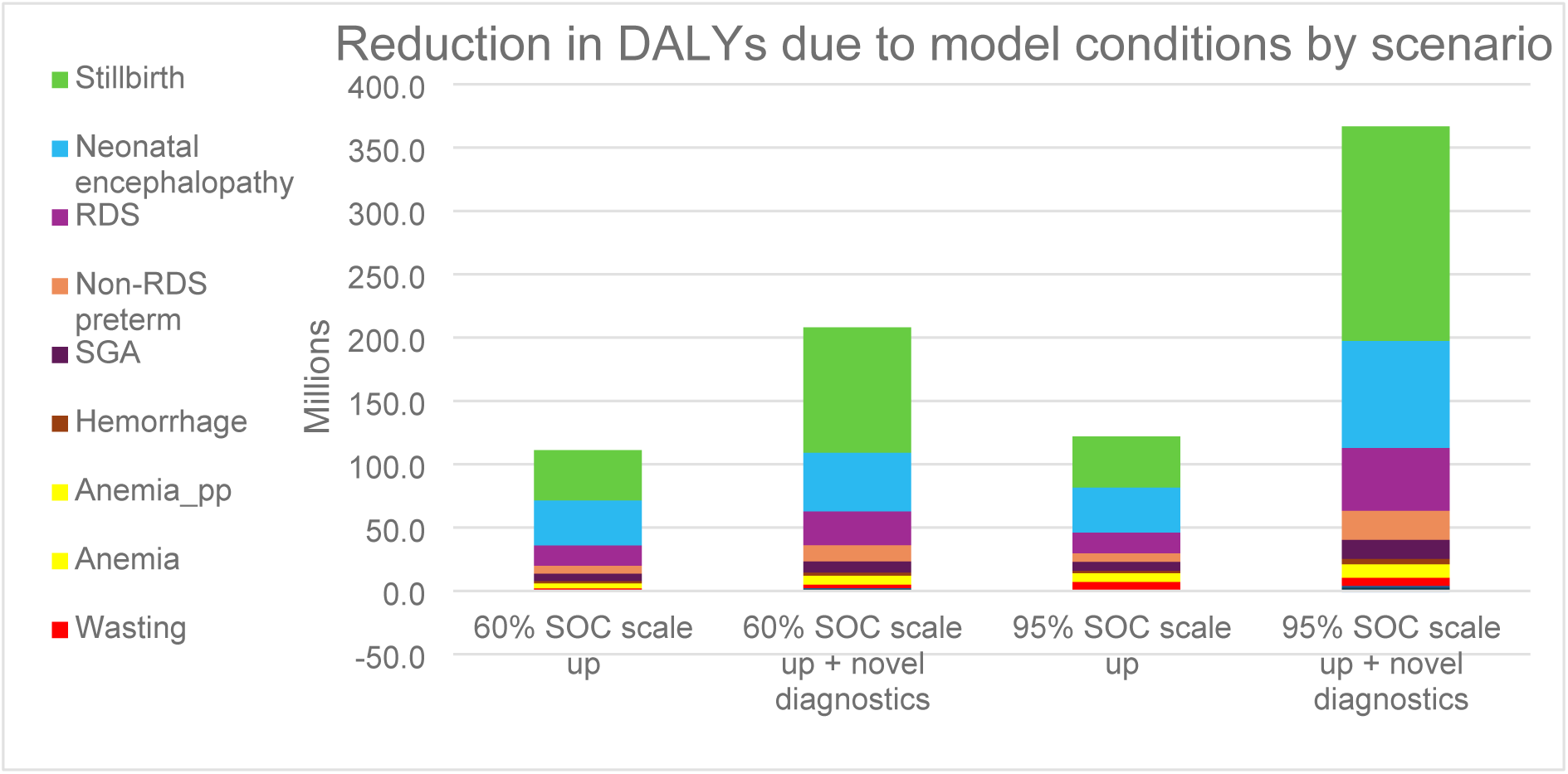
Reduction in DALYs by condition and scenario. Note: a negative value appears on the y-axis due to an increase of DALYs associated with infant conditions (wasting) relative to baseline, since the total number of pregnancies continuing to infancy is increased due to reductions in maternal and neonatal mortalities. Since no interventions directly target infant conditions the total number of morbidities and mortalities (and therefore DALYs) associated with the conditions is increased.

Including novel diagnostic products in the scale up scenario had a considerable impact on mortalities and stillbirths averted compared to the baseline scenario (Figure 3). Scaling up all SOC and novel interventions to 60% or 95% coverage averted an additional **207 million and 365 million DALYs** compared to the baseline, respectively (Table 3, Figure 2). In particular, including novel diagnostics at 60% coverage had a greater impact than the 95% coverage SOC scale up scenario.

**Figure 3.**
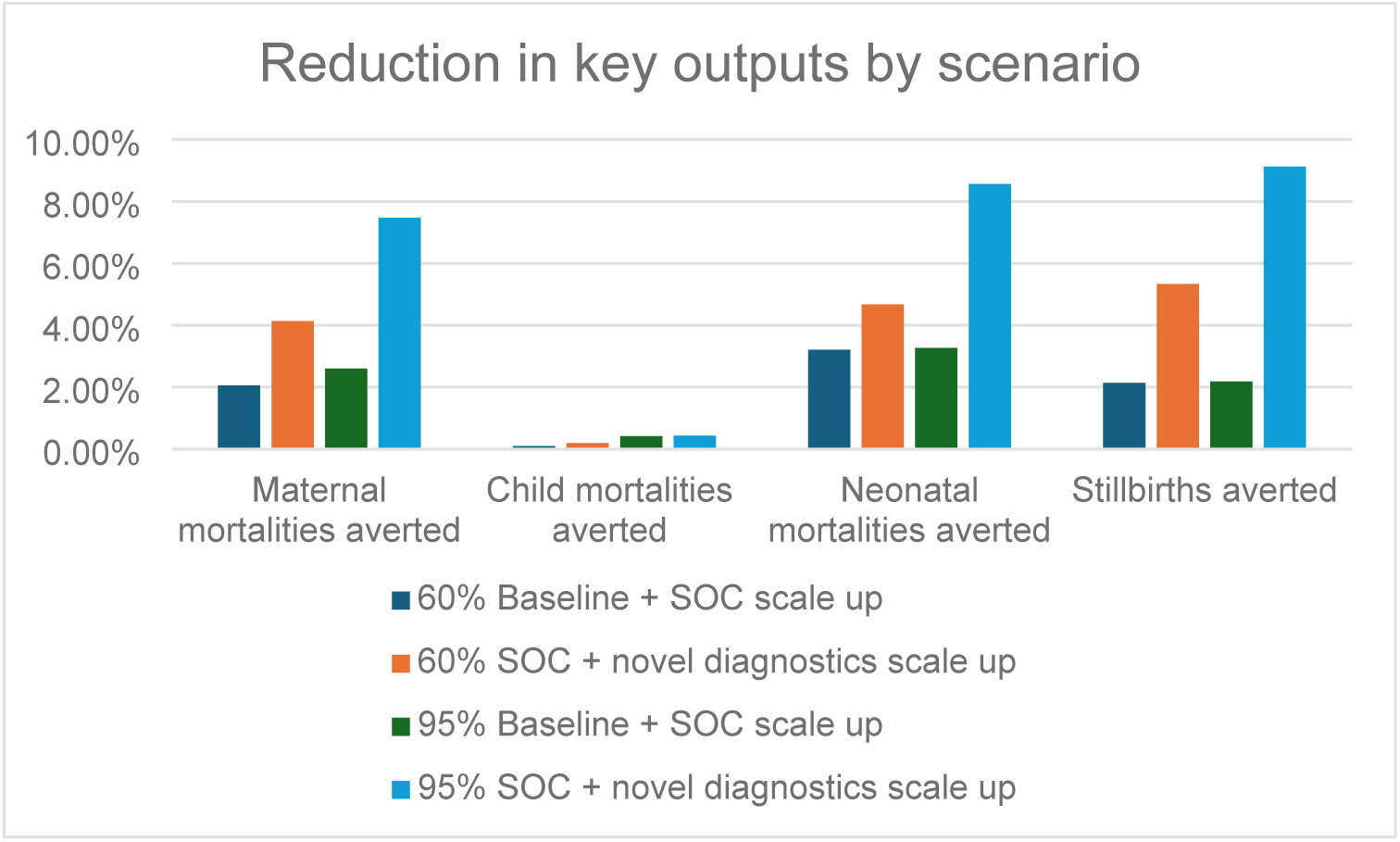
Reduction in key model outcomes for scale up scenario compared to baseline 2024-2040

For the set of interventions considered in this example, the conditions with the greatest reduction in DALYs were stillbirths, wasting and birth asphyxia. The majority of the DALY reductions for these conditions were due to scaling up AI ultrasound, preeclampsia screening, SOC RUTF and SOC ultrasound.

### Reduction in mortalities/stillbirths by scenario

## Discussion

We have created a model that captures dynamic interactions between MNCH conditions, including across trimesters, to account for the full impact of early detection and prevention of conditions during pregnancy. In addition, the model quantifies direct and indirect health benefits and allows for flexible integration of new interventions to enable the assessment of products under development.

A use case has been presented as an example. The example analysis shows there is potential to avert a large number of DALYs through greater coverage of existing evidence-based interventions. If the coverage of SOC interventions in LMICs could be increased to 60%, 111 million DALYs could be averted over 2024-2040 compared to the SOC scenario. This increased to 121 million DALYs averted if intervention coverage were increased to 95% by 2030, with diminishing returns due to ANC coverage restricting the widespread delivery of some interventions. Investing in increasing the coverage of SOC interventions could have a significant impact on MNCH outcomes.

Additionally, the analysis shows that investment in novel diagnostic interventions can have a significant impact on the reduction of DALYs. The scenario with SOC and novel diagnostics scaled to 60% had a bigger impact on key outputs than scaling SOC interventions to an optimistic 95%. If both SOC interventions and novel diagnostic products were increased to 60% coverage in LMICs, 207 million DALYs could be averted over 2024-2040 compared to the baseline scenario. Scaling up the entire package of interventions to an aspirational 95% produced additional benefits. Of the novel interventions considered, AI ultrasound had the largest modelled impact, through the possibility of increased accessibility compared to the SOC ultrasound. When scaled up to 95% coverage in isolation, AI ultrasound averted 248 million DALYs in LMICs over 2024-2040 which was 68% of the DALYs averted in the SOC + novel diagnostics scale up with 95% coverage scenario. These results showed that even at low coverage levels investing in novel interventions could be more beneficial than focusing on increasing the coverage levels on current interventions. In order to make progress towards the SDG targets funds need to be invested in the highest yield interventions. This analysis also highlights why it is critical to capture downstream impacts, since a majority of the benefits were from indirect DALYs averted. This model can therefore help guide clinical decisions surrounding long-term and broader health consequences of treatment. Future studies may include a bigger set downstream impacts to capture the full set of benefits arising from a single interventions.

An earlier version of this model was used to produce outcomes as part of a recent report for the BMGF which highlighted maternal and neonatal mortality worldwide(12). The report notes that progress has stalled and offers insights into innovations needed to improve maternal, neonatal and childhood health and that reaching the SDG targets 3.1 and 3.2 will require significant improvement and investment in interventions with the highest impact. The report analysed how novel interventions might fast-track progress towards these targets. Modelling realistic intervention coverage rollout targets can identify where the largest impact and potential improvements can be, which can be used to support investment decisions. We hope that the model described in this paper will aid in decision making surrounding MNCH interventions and therefore help reduce the global burden of maternal, neonatal and child health conditions as policy makers aim to reach these targets.

The model used in this study has some similarities and differences to other models in the field. Some other tools that have been used to answer questions surrounding MNCH are the Lives Saved Tool (LiST)(3), Equitable Impact Sensitive Tool (EQUIST)(20), One Health Tool, Optima(21), Maternal and Newborn Directed Assessment of Technology (MANDATE)(22), Health Equity Assessment Toolkit (HEAT and HEAT Plus)(23) and REDUCE-ALIVE(21). Depending on the country context and key questions of decision-makers, one tool may be more appropriate to use than another. In particular, LiST has been successfully used in a number of country studies to support funding bodies and policy makers in decisions surrounding the impact of WHO recommended interventions(24). LiST is used to scale up key maternal and child health interventions in order to model health impacts and cost scenarios. The model presented in this paper shares a number of similarities with LiST. In both models, modifying coverages of interventions produces estimates of maternal and child (up to 5 years) deaths, mortality rates/ratios, deaths averted, intermediate outcomes, and single- and multiple-country scenarios. Intervention coverages can be projected into the future in order to model a number of different scenarios. However, they contrast in the way they calculate health outcomes in populations that are not directly targeted by a particular intervention. EQUIST, HEAT and HEAT-PLUS focus on including equity into policy and coverage decisions related to MNCH. Currently the model developed in this paper does not consider equity in its analysis. This is a possible direction for the future.

A unique strength of this model is the ability to flexibly implement new interventions. This allows a user to model the potential impact of novel interventions as well as a wide range of current interventions without requiring external developers to update the model. In cases where there is limited funding for research and development it is critical to understand the capacity for novel interventions and interventions that have not yet made it to market to impact the entire MNCH area. A further advantage of this model is that it includes trimester timesteps. This means that we can more accurately model early detection and treatment of conditions throughout pregnancy and the associated benefits. This can have further effects on downstream conditions in the maternal, neonatal and child populations. Additionally, as discussed, the model is set up to allow interactions between conditions. In such a way the complex interaction and downstream impacts of conditions may be used to test potential impact pathways. This may be advantageous to policy makers who are interested in the way conditions and packages of interventions interact.

As an example, AI ultrasound is a novel intervention which has a wider coverage than traditional ultrasound due its ability to be used by a larger number of health care workers. Due to the model structure we were able to integrate this novel intervention to better understand its potential impacts. Although AI ultrasound only targets pregnant women in the ANC module, it has the majority of its impacts downstream. This is due to treatment of conditions which are detected through AI ultrasound. Furthermore, trimester timesteps within the model allow for early AI ultrasound to detect and indicate early treatment for conditions during pregnancy, which reduce adverse outcomes during childbirth, in neonates and in children under 5 years.

There are some limitations to this study. Firstly, in the theoretical intervention coverage scenarios, implementation challenges were not considered, including the adequacy of healthcare workforces and infrastructure to scale up interventions. Secondly, analyses were performed on large aggregated LMIC regions and so may not capture the intricacies of health systems related to individual countries. Country-specific analyses would be warranted with a broader set of interventions to inform policy. Thirdly, this study does not include consideration of costs or cost-effectiveness, which could be the subject of further analyses. Fourthly, although it would be useful to validate the outcomes of this use case against a similar analysis in existing models such as LiST, this is not possible due to the consideration of novel interventions. Similarly, we cannot properly identify the extent to which indirect effects of interventions are underestimated, since in reality we cannot easily identify which impacts are due to upstream impacts. Some future work may involve modelling the extent to which these indirect effects are under calculated. Additional future work may involve using the underlying Atomica software package to produce allocative efficiency analyses in-order-to aid decisions surrounding which interventions to prioritise.

## Conclusion

We have presented a mathematical modeling tool which can project the direct and indirect impact of packages of existing and novel interventions on a range of MNCH outcomes. The model can be flexibly adapted to include new interventions, and can be applied at a global, regional, national or sub-national level. Our example analysis identified that there is significant opportunity to avert morbidities, mortalities and DALYs in LMICs by expanding access to SOC and novel MNCH interventions.

## Data Availability

The data supporting the findings of this study are provided in the supplementary material. These data were compiled from source datasets, with citations and access details provided in the supplementary files. Further inquiries can be directed to the corresponding author.

## Acknowledgements

We gratefully acknowledge our Gates Foundation colleagues Chris Troeger, Laura Lamberti and Sarah Metzger, for their contributions to the design of this study, the collection of data and their insightful feedback and perspectives on this manuscript.

